# Family Vulnerability Scale: evidence of content and internal structure validity

**DOI:** 10.1101/2023.01.12.23284419

**Authors:** Evelyn Lima de Souza, Ilana Eshriqui, Flávio Rebustini, Eliana Tiemi Masuda, Francisco Timbó de Paiva Neto, Ricardo Macedo Lima, Daiana Bonfim

## Abstract

**Introduction:** territory view based on families’ vulnerability strata allows identifying different health needs that, in their turn, can guide healthcare at primary care scope. Although there are instruments aimed at measuring family vulnerability, they still need robust validity evidences; therefore, they represent a limitation for usability in a country showing multiple socioeconomic and cultural realities, such as Brazil. The present study introduces the development and search for evidences about the validity of the Family Vulnerability Scale for Brazil, known as EVFAM-BR.

**Methods:** items were generated through exploratory qualitative study carried out with 123 professionals. Collected data subsidized the generation of 92 initial items that were subjected to a panel of multi-regional and multi-disciplinary judges (n = 73) to calculate the Content Validity Ratio (CVR) – this process resulted in a version of the scale comprising 38 items. Subsequently, it was applied to 1,255 individuals to find evidences about the internal-structure validity by using the Exploratory Factor Analysis (EFA). Dimensionality was assessed through Robust Parallel Analysis and the model was tested through cross-validation to find EVFAM-BR’s final version.

**Results:** the final version comprised 14 items distributed into four domains, with explained variance of 79.02%. All indicators were within adequate and satisfactory limits, without any cross-loading and Heywood Case issues. Reliability indices also reached adequate levels (_α_ = 0.71; _ω_ = 0.70; glb = 0.83 and ORION ranging from 0.80 to 0.93, between domains). Instrument’s score was subjected to normalization, and it pointed towards three vulnerability strata (0 to 4 – Low; 5 to 6 – Moderate; 7 to 14 - High).

**Conclusion:** the scale showed satisfactory validity evidences, which were consistent, reliable ad robust; it led to a synthesized instrument capable of accurately measuring and differentiating family vulnerability in the primary care territory, in Brazil.

## Introduction

The term vulnerability is the core of debates in different knowledge fields; etymologically it would come from Latin “vulnerare” (harm, impair) and “bile” (susceptible) [1]. However, the understanding about vulnerability shows variations when different dimensions linked to this topic are taken into account [2]. Vulnerability, in the Bioethics field, refers to being in danger or exposed to risk due to individual weakness, which is a feature inherent to human beings [3]. As for the healthcare field, this term has a broader meaning; it is associated with acknowledging humans as susceptible to damage or to risks within the health/illness process due to social disadvantages [4].

Although the literature presents different vulnerability definitions and assesses individual predictors, the concept of family vulnerability is measurable through different ways [4] because it must take into consideration this phenomenon from multiple aspects that, in their turn, are linked to health needs of members from a given family. Aspects related to the health condition of members composing the family nucleus, as well as the community and social context, are elements to be taken into account at the time to investigate vulnerability in families [5,6].

Thus, health services and managers must consider family vulnerability to organize healthcare practices, mainly from the population perspective [7]. Accordingly, territory view from vulnerability strata perspectives subsidizes the process to identity health needs in different population groups [2]. Besides, family vulnerability stratification is essential at the time to plan the offer of services in a given territory, since it would help achieving equity and qualified of population-based care management. Hence, considering health teams activities and the care provision order in the Health Care Network based on the needs identified through family vulnerability demands, it is needed to deepening in aspects composing family vulnerability.

Instruments and initiatives to ensure the observation of aspects likely presented by family vulnerability as components to labor process organizations remain scarce. Some global experiences are linked to this phenomenon: vulnerability-measuring background lies on the United Nations Program for Development (UNPD); back in 2004, it elaborated the Disaster Risk Index (DRI) to measure and compare countries within a process based on physical, social, economic and environmental factors [8]. Subsequently, in 2006, the Autonomous University of Madrid (Spain) estimated the degree of vulnerability of citizens assumingly susceptible to lack of social protection, based on countries belonging to the Organization for Economic Cooperation and Development (OECD) [9]. With respect to the Latin scene, one research aimed at measuring family vulnerability rates in a Colombian municipality based on a sample of families from all socioeconomic strata living in an urban zone and in a rural one [10].

Other initiatives have been introduced and they aimed at developing instruments to be used by primary healthcare (PHC) teams to measure family vulnerability and, consequently, to contribute to plan healthcare provision in the Brazilian territory [11-15]. However, instruments so far developed and used for such a purpose still need robust evidences of their validity, since they present limitations to be used in a country with continental dimensions, and multiple socioeconomic and cultural realities, like Brazil.

Thus, it is essential reasoning about the concept of family vulnerability, with emphasis on a diversified population and from this perspective, to develop an instrument which could be used in a standardized way in national scope. Then, the present study aim to seek validity evidences about the content and internal structure of the Family Vulnerability Scale for Brazil (EVFAM-BR).

## Materials and methods

The present study followed a psychometric nature design to seek evidences about both content validity (stage 1) and internal structure (stage 2) based on current recommendations by the Educational Research Association (AERA), the American Psychological Association (APA) and the National Council on Measurement in Education (NCME) [16]. The study was approved by the Ethics Research Committee of *Hospital Israelita Albert Einstein*, which was approved on October 22, 2019 (nº 3.674.106, CAAE 12395919.0.0000.0071).

### Stage 1: content validity evidences

PHC professionals from all Brazilian geographic regions were invited to join the first qualitative exploratory stage of the study to define the concept of “family vulnerability” and to identify factors likely associated with it, in order to subsidize the development of items for the instrument. It was done to identify different understandings about family vulnerability in different geographic regions countrywide.

The invitation was made based on the snowball method [17], by WhatsApp and e-mail. Using the RedCap® electronic tool [18,19], an online semi-structured questionnaire was made available for participants after they read the free consent form and formally accepted to join the study.

The questionnaire comprised (i) respondents’ socioeconomic, demographic and health profile identification, (ii) open questions about the concept of vulnerability and scale applicability, and (iii) multiple-choice questions about the relevance of measuring family vulnerability; the questions were distributed into 46 items elaborated from individual and domestic registration forms used at the Brazilian public healthcare system through e-SUS PHC system [20,21]. Frequencies of responses for items expressed in multi-choice questions and the group of aspects identified in open questions were taken into consideration based on the Content Analysis Technique in order to elaborate the first version of items [22] – they were developed in an interrogative way to allow dichotomous answers (“0 – no” and “1 – yes”).

Items developed from the previous stage were subjected to an extensive panel of multi-regional and multi-disciplinary judges. The panel encompassed health professionals, scholars and psychometrists who were invited to join the study through the snowball method.

The large number of judges was explained by the need of calculating and applying the instrument at national scope. The judges judged the items in the first version of the instrument based on relevance and clarity criteria, as well as were enquired about the need of changing the writing of any item. Then, option was made to apply the Content Validity Ratio (CVR) scale [23] as the validity index to select the items. CVR is calculated based on the number of judges in the panel [24,25] in order to allow the adoption of a larger number of judges. CVR was initially applied to assess the relevance of a given item in order to check whether it effectively measures the latent variable: family vulnerability. CVR was represented as CVR-1 (the Item’s CVR) and CVR-E (Scale CVR) – this last one corresponds to mean recorded for the CVR criteria. It is important highlighting that a modified CVR version with two points, namely “no” and “yes”, was adopted. The original version had three points and did not have an effective practical effect, since, for CVR calculation purposes, it used to become dichotomous. This procedure was already adopted in studies [26,27].

Oftentimes, the mean recorded for the assessed questionnaires is adopted; therefore, a hierarchic flow was herein adopted, since other indicators, such as item’s clarity, were only analyzed after judges showed its relevance to testify that the item actually assesses the instrument’s latent variable. The application of requirements in the same stage tends to inflate mean CVR and to launch items that do not measure the latent variable to the next stage. Accordingly, as pointed out by DeVellis [28], a given item can be relevant, but its words might be problematic. Thus, the second stage refers to items’ clarity (whether the item is well written in terms of its semantics). The third stage assessed the need of changing the items’ writing. The mean recorded for CVR was only applied to items that adhered to the phenomenon.

### Stage 2: Evidences about internal structure validity

The version subjected to content evidences was applied to users of PHC services to find evidences about internal structure validity.

All data collectors were previously trained and clarified about the informed consent form application, as well as about research aim, methodology and questions. Study presentation and data collection flow were also previously carried out with teams from the participating PHC services.

PHC services selection was based on municipalities presenting the largest population of attendees of the Program to Support the Institutional Development by the Unified Healthcare System (Proadi-SUS) in Brazil, PlanificaSUS [29]. It was done by including at least one PHC service from each geographic region in the country. Thus, data collection was carried out in 11 PHC services: 1 in Northern Brazil (Roraima State), 1 in the Northeastern region (Pernambuco State) and 2 in Midwestern Brazil (Mato Grosso State), 5 in the Southeastern region (São Paulo and Minas Gerais states) and 2 in Southern Brazil (Paraná State).

The Covid-19 pandemic met the first data collection stage (from June to November 2020); therefore, interviews with São Paulo healthcare unit users were carried out by phone. Service managers were aware of it, since they provided information to identify interviewees in the territory. As for the second data collection time (from May to August 2022), users who had attended the participating PHC service at data-collection day were asked to join the study.

Over 18-year-old participants were informed about the informed consent form at both data-collection times and they only joined the research after signing it. Subsequently, the structured questionnaire about the family vulnerability scale was applied, and it was followed by participants featuring in RedCap® [18,19].

### Statistical analysis

#### Exploratory factor analysis

The first stage of the analysis aimed at assessing whether the collected data were prone to factorial through Measure of Sampling Adequacy (MSA). Bartlett sphericity, determinant of the matrix and Kaiser-Meyer-Olkin (KMO) were assessed at this stage. Besides assessing the dataset items, individual analysis was also assessed, as recommended by Lorenzo-Seva and Ferrando [30]. The inadequacy of items to be factored can affect model solution. Missing data were treated through the multiple imputation technique [31].

Dimensionality testing was carried out through Parallel Analysis, based on Optimal implementation of Parallel Analysis (PA) and Minimun rank factor analysis to minimize the common variance of residues [32]. PA was implemented through permutation with 500 random matrices. Dimensionality in exploratory factorial analysis (unrestricted model) was tested through Parallel Analysis, which has been considered more robust and accurate to test it [33-37].

Tetrachoric matrix estimates were carried out through Bayes Modal Estimation [38], with Smoothing Ridge [39]. The use of tetrachoric/polychoric correlations tends to increase the model’s accuracy in comparison to Pearson’s correlation [40,41].

Factors’ extraction was performed through the RULS technique (Robust Unweighted Least Squares), which reduces the residues in matrices that are more robust in terms of abnormal data [42]. Promin oblique rotation would be used in case the instrument emerged as multi-dimensional [43].

UNICO (Unidimensional Congruence > 0.95), ECV (Explained Common Variance > 0.80 – Quinn, 2014) and MIREAL (Mean of Item Residual Absolute Loadings < 0.30) were adopted as unidimensionality assessment indicator [44].

#### Quality parameters of the instrument

Instrument explained variance must be close to 60% [45]. Initial factorial load of 0.30 is recommended when the sample comprises less than 300 individuals [45]; communities must present values higher than 0.40 [46]. The maintenance or removal of a given model item depend on factorial load magnitude, on the communities and on the existence of cross-loading and Heywood cases, as well as on the impermeability of factors. The unique directional correlation (Eta) through Pratt’s Measure was adopted to increase the accuracy of decision-making about the maintenance or removal of a given item [47,48].

#### Reliability

Reliability was measured through four indicators: Cronbach’s alpha [49], Greatest Lower Bound – glb [50], Omega [51] - all three by means of Bayesian estimates - and ORION (Overall Reliability of Fully-Informative prior Oblique N-EAP scores) [52].

Cross-validation was applied to increase the model’s reliability and replicability; the Houdolt technique was also herein applied [53]. This technique divides the dataset into a training sample - that can range from 10%, 30% to 50% - and into a dataset known as test dataset [53]. The dataset in the present study was split in half by randomly choosing the items. The Solomon technique [54] was adopted, so that dataset division could be random and respect factorability’s equivalence. The datasets were labeled as follows: Full Sample (FS; n = 1,255); Training Sample (TrS n = 627) and Test Sample (TsS; n = 628). According to Brown [55], cross-validation can be carried out either through EFA or Confirmatory Factor Analysis (CFA). FS analysis will only take place if the model found in TrS and TsS can be replicated. This procedure was already adopted in previous studies [56,57], and it follows contemporary recommendations [58].

#### Descriptive study and standardization

An exploratory descriptive study of general scores recorded for the Family Vulnerability Scale (FVS - EVFAM-BR, in Brazilian Portuguese) was carried out after a solution for the internal structure was found. Results recorded for the items and for total score were represented by answers’ frequency, median (Md), interquartile interval (IIQ), amplitude (amp), minimum (min) and maximun (max) value.

Standardization, in the first stage, was performed by identifying score cuts based on participants’ distribution. Despite this process, although participants’ distribution is recurrent in standardization studies, it can present distortions, because the score is not directly analyzed, but it can be taken as consequence of participants’ position in the cutting points. Discriminant analysis of each one of the limits and Family Vulnerability Scale scores were used to improve the accuracy of proposed cuts (within the limit) and to assess the predictive ability to classify the individuals. The discriminant analysis aims at better understanding group differences and at predicting the probability of an entity (individual or object) to perceive a specific class or group, based on several independent variables of the metrics [59]. Boedeker and Kearns [60] identified a better performance by the discriminating analysis in comparison to many other techniques applied for the same purpose. Besides, it allows determining the independent variable mostly accounting for differences in mean score profiles in two or more groups [59]. Tabachnick and Fidell [61] added to this information by stating that the aim of the discriminating analysis is to predict the group’s participation based on a set of predictors. Accordingly, it is possible confirming whether the groups formed from the distribution process have properly classified individuals within the established limits.

Data were analyzed in statistic software Factor 12.01.01, SPSS v.23 and JASP 16.04.

## Results

### Content validity evidences

In total, 123 professionals from the five Brazilian regions joined the first stage of the research to define the concept of “family vulnerability”: 48.8% of them came from Northeastern Brazil; 21.1% from Southeastern Brazil; 17.9% from Southern Brazil, 8.9% from Northern Brazil and 3.3% from Midwestern Brazil. Most professionals belonged to the female sex (82.9%), approximately 40% of them had at least 10-year experience in PHC and 48.8% reported to have specialization degree and higher schooling profiles. Nurses were the professional category accounting for the highest participation in this stage (48.3%); they were followed by Community Health Agents – CHA – (10.7%). The first version of the instrument counted on 92 items; it was developed from factors’ responses that, at first, could be associated with the concept of family vulnerability.

A panel of multi-regional and multi-disciplinary judges was set for the second stage; it aimed at identifying content validity evidences. This panel comprised 73 judges: 61.7% from Southeastern Brazil, 15.1% from Southern Brazil, 9.6% Northeastern Brazil, 6.8% from Northern Brazil and 6.8% from Midwestern Brazil. Most of them belonged to the female sex (79.5%), more than half of them had less than 10-year experience in PHC (57.5%) and specialization as higher schooling profile (51.4%). Nurses were the professional category accounting for the highest participation in the panel (50.7%), they were followed by physicians (16.55). CVR was applied to judges’ answers. CVR critical value was established at CVR > 0.12, which was defined based on the participation of 73 judges.

CVR calculation led to the exclusion of 54 items, and it resulted in scale version comprising 38 items linked to socioeconomic and demographic aspects, access to healthcare services, health condition and life style. In order to achieve a better understanding of it, 12 of the 38 items were rewritten based on recommendations from the panel of judges. It must be clear that only items 2 and 15 (Table 1) recorded CVR lower than the critical value; therefore, their text was revised. The other 10 items did not suffer any change; writing adjustments were made in the original text just to meet CVR’s critical value. The version presenting evidence of content validity (38 items) was taken into consideration in the stage to evidence internal structure validity.

**Table 1.** Items that remained in the scale after Content Validity Ratio (CVR) application.

| Item | Content Validity Ratio (CVR) |  |  |  |
| --- | --- | --- | --- | --- |
|  | Relevance | Clarity | Need of change<sup>a</sup> | New text of the item |
| 1. Is there lack of basic sanitation system in the neighborhood you live in? | 0.12 | 0.12 | 0.78 | Is there open sewer in your neighborhood? |
| 2. Do you drink untreated water in your house? | 0.12 | 0.07 | 0.86 | Does the water in your house lack treatment? |
| 3. Does your house face the risk of flood? | 0.18 | 0.40 | 1.00 | - |
| 4. Do you live close to drug dealing areas? | 0.12 | 0.40 | 0.95 | - |
| 5. Does anyone at your house live close to violent people? | 0.21 | 0.32 | 0.97 | - |
| 6. Has anyone in your house been victim of violence? | 0.26 | 0.34 | 1.00 | - |
| 7. Is there violence in your house? | 0.23 | 0.32 | 1.00 | - |
| 8. Is anyone in your house in legal custody condition? | 0.12 | 0.26 | 0.86 | Is anyone in your Family in jail? |
| 9. Is anyone in your house facing financial issues? | 0.15 | 0.12 | 0.97 | - |
| 10. Does anyone lack money to fulfill household needs ? | 0.12 | 0.21 | 0.97 | - |
| 11. Does anyone in your house is a *Bolsa Família* beneficiary? | 0.12 | 0.32 | 1.00 | - |
| 12. Is anyone in your house a BPC (continued benefit)/LOAS (Social Security Organic Law) beneficiary? | 0.15 | 0.21 | 0.86 | Does anyone in your house get health benefit (BPC /LOAS)? |
| 13. Have any health professional ever mentioned that someone in your house presents obesity? | 0.12 | 0.15 | 0.86 | - |
| 14. Have any health professional ever mentioned that someone in your house suffers with malnutrition? | 0.15 | 0.12 | 0.89 | - |
| 15. Is there any difficulty in making sure about food variety in your house? | 0.15 | 0.10 | 0.92 | It is hard to make sure about the access to different food types? |
| 16. Does anyone in your house starve? | 0.21 | 0.26 | 0.92 | - |
| 17. Does anyone in your house have drug addiction? | 0.23 | 0.26 | 0.89 | Does anyone in your house use illegal drugs? |
| 18. Is anyone in your house an alcohol abuser? | 0.23 | 0.26 | 0.92 | - |
| 19. Does anyone in your house use controlled medication? | 0.15 | 0.37 | 0.95 | - |
| 20. Does anyone in your house use 5, or more, medications a day? | 0.18 | 0.40 | 0.95 | Does anyone in your house uses 5, or more, medications on a daily basis? |
| 21. Does anyone in your house have a health condition that demands long-term caregiving? | 0.21 | 0.26 | 0.97 | Does anyone in your house have a health condition that requires continuous care? |
| 22. Is anyone in your house impaired to perform daily activities? | 0.18 | 0.23 | 0.86 | - |
| 23. Is anyone in your house helped by others to accomplish its own daily healthcare procedures? | 0.15 | 0.21 | 0.95 | Does anyone in your house need help to accomplish its own daily healthcare procedures? |
| 24. Does anyone in your house present any disability? | 0.12 | 0.37 | 1.00 | - |
| 25. Does anyone in your house have any intellectual/cognitive disability? | 0.15 | 0.29 | 0.84 | - |
| 26. Does anyone in your house have mental issues? | 0.21 | 0.34 | 1.00 | - |
| 27. Does anyone in your house have HIV/aids? | 0.15 | 0.32 | 0.97 | - |
| 28. Is anyone in your house sick in bed? | 0.15 | 0.32 | 1.00 | - |
| 29. Does anyone in your house often go to urgency and emergency units? | 0.18 | 0.29 | 0.95 | - |
| 30. Does anyone in your house do not know the UBS/healthcare unit team in charge of your family? | 0.15 | 0.32 | 0.95 | - |
| 31. Did anyone in your house have a child without wanting it? | 0.15 | 0.29 | 0.95 | Has anyone in your house had an unplanned child? |
| 32. Did anyone in your house have a child before turning 20 years old? | 0.12 | 0.37 | 0.97 | - |
| 33. Has anyone in your house had its mother absent in childhood? | 0.18 | 0.34 | 1.00 | - |
| 34. Has anyone in your house had an absent father in the childhood? | 0.12 | 0.32 | 1.00 | - |
| 35. Has anyone in your house faced abandonment by the family? | 0.21 | 0.18 | 0.95 | - |
| 36. Are children in your house out of school? | 0.12 | 0.32 | 0.92 | Are there children in your house out of school? |
| 37. Are there adolescents in your hose out of school? | 0.12 | 0.37 | 0.97 | Do you have any adolescent in your house out of school? |
| 38. Are there under 14-year-old individuals in your house that have a job? | 0.21 | 0.23 | 0.95 | - |
| **Mean CVR** | **0.16** | **0.27** | **0.94** | - |

<sup>a</sup> Accordingly, CVR values higher than the critical value point towards no need of changing the writings, although some items under this condition were rewritten in order to improve semantics' adequacy.

### Evidences about internal structure validity

In total, 1,584 users who attended the 11 PHC services during data collection were invited to join this research stage. However, only 1,505 (95%) of them accepted the invitation and signed the informed consent form. Only 1,255 of them completed the interview for the application of the scale version presenting content validity evidences. This sample represented the study’s final sample and this version presented content validity evidences. Table 2 introduces participants’ description of this study stage.

**Table 2.** Participants’ featuring.

| Variables (n=1255) | Categories | N (%) |
| --- | --- | --- |
| Age in years<sup>a</sup> | - | 43.3 (15.5%) |
| Sex (n=756) | Female | 551 (43.9%) |
|  | Male | 205 (16.3%) |
| Race/skin color (n=1217) | White | 386 (30.8%) |
|  | Brown | 640 (51.0%) |
|  | Black | 150 (12.0%) |
|  | Yellow | 23 (1.8%) |
|  | Indigenous | 18 (1.4%) |
| Schooling (in years) (n=1237) | 0 to 4 years | 160 (12.7%) |
|  | 5 to 8 years | 225 (17.9%) |
|  | 9 to 11 years | 247 (19.7%) |
|  | 12 to 15 years | 480 (38.2%) |
|  | Over 16 years | 125 (10.0%) |
| Job (n=1237) | Unemployed or does not have a job | 447 (35.6%) |
|  | Employer | 1 (0.1%) |
|  | Self-employed without social security | 111 (8.8%) |
|  | Self-employed with social security | 47 (3.7%) |
|  | Wage owner | 413 (32.9%) |
|  | retired/pensioner | 181 (14.4%) |
|  | Others | 37 (2.9%) |
| income (in minimum wage)<sup>b</sup> (n=726) | No income | 58 (4.6%) |
|  | Up to 1 minimum wage | 327 (26.1%) |
|  | > 1 and lower than 2 minimum wages | 179 (14.3%) |
|  | >= 2 and lower than 4 minimum wages | 115 (9.2%) |
|  | >= 4 lower than 10 minimum wages | 31 (2.5%) |
|  | >= 10 minimum wages | 4 (0.3%) |
|  | Does not know | 12 (1.0%) |
| Number of children<br>(n=759) | 1 child | 140 (11.2%) |
|  | 2 children | 198 (15.8%) |
|  | 3 children | 129 (10.3%) |
|  | More than 3 children | 129 (10.3%) |
|  | Expecting the first child | 19 (1.5%) |
|  | Does not have children | 144 (11.5%) |
| Number of households<br>per room in the house<br>(n=1223) | <1 | 406 (32.4%) |
|  | 1 | 697 (55.5%) |
|  | >1 | 120 (9.6%) |
| Private healthcare<br>insurance (n=1231) | Yes | 180 (14.3%) |
|  | No | 1,051 (83.7%) |
| Systemic High Blood<br>Pressure (n=1240) | Yes | 212 (16.9%) |
|  | No | 1,028 (81.9%) |
| Diabetes Mellitus<br>(n=1238) | Yes | 96 (7.6%) |
|  | No | 1,142 (91%) |
| Cancer (current)<br>(n=1237) | Yes | 6 (0.5%) |
|  | No | 1,231 (98.1%) |
| Heart disease<br>(n=1240) | Yes | 72 (5.7%) |
|  | No | 1,168 (93.1%) |
| Intellectual/cognitive<br>impairment (n=812) | Yes | 15 (1.2%) |
|  | No | 797 (63.5%) |
| Tuberculosis (n=1240) | Yes | 3 (0.2%) |
|  | No | 1,237 (98.6%) |
| Leprosy (n=1239) | Yes | 1 (0.1%) |
|  | No | 1,238 (98.6%) |
| Kidney issues<br>(n=1237) | Yes | 35 (2.8%) |
|  | No | 1,202 (95.8%) |
| Breathing issues<br>(n=1232) | Yes | 96 (7.6%) |
|  | No | 1,136 (90.5%) |
| Mental issues<br>diagnosis (n=1238) | Yes | 30 (2.4%) |
|  | No | 1,208 (96.3%) |

<sup>a</sup> continuous numerical variable described as mean and standard deviation.
<sup>b</sup> one minimum wage corresponds to R\$1,212.00 (in Brazilian Real, in 2022).

#### Factorability

The evaluation of sample adequacy measures is the first step of the factorability analysis; it aims at assessing dataset factorability and whether factorial analyses are applicable. Dataset’s general data have shown good factorability: Fs recorded KMO (0.75), Bartlett Sphericity = 6,084.6 (df = 91; P < 0.0001) and determinant of the matrix = 0.00001. As for TrS: KMO (0.74), Bartlett Sphericity = 6,153.7 (df = 91; P < 0.0001) and determinant of the matrix = 0.00001; TsS: KMO (0.71), Bartlett Sphericity = 2,617.3 (df = 91; P < 0.0001) and determinant of the matrix = 0.0002. Although general indices presented good indicators, 4 of the 38 initial items have shown factorability issues in three datasets (27 - Does anyone in your house have HIV/aids?; 36 - Are there children in your house out of school?; 37 - Do you have any adolescent in your house out of school?; and 38 - Are there under 14-year-old individuals in your house that have a job?). They were excluded from the analyses based on recommendations by Lorenzo-Seva and Ferrando [30].

#### Dimensionability

The first analyses were carried out in TrS. Dimensionality analyzed through parallel analysis pointed towards a 4-dimension model. Closeness of dimensionality values kept the indication for multi-dimensional model: Single = 0.82; ECV = 0.65 and MIREAL = 0.37. Thirteen (13) of the 34 items forming the initial analysis did not present substantial factorial load in the model. Accordingly, the process to remove items in order to adjust the model followed two principles: quantitative (statistical adjustment) and qualitative (interpretability) adjustment. The choice for removing an item was carried out by taking into consideration the set of primary indicators: factorial load, communality, Eta of Pratt’s Importance Measure, existence of cross-loading, Heywood case and model adjustment indices. The items were removed from the scale up to the time the two principles were congruent to each other, and it resulted in a model for 4-dimension TrS, with 14 items with proper statistical adjustment, open for interpretability. The parallel analysis kept on pointing out a 4-dimension solution and explained variance of 78.66%. This model was replicated in TsS and FS. Both datasets confirmed the 4-dimension model; furthermore, the closeness of dimensionality values reinforced the multi-dimensional model (Table 3) for the three datasets.

**Table 3.** Items’ I-Unico, I-ECV and I-Real values.

| Item | I-UNICO |  |  | I-ECV |  |  | I-REAL |  |  |
| --- | --- | --- | --- | --- | --- | --- | --- | --- | --- |
|  | TrS | TsS | FS | TrS | TsS | FS | TrS | TsS | FS |
| 1. Is anyone in your house facing financial issues? | 1.000 | 0.958 | 1.000 | 0.991 | 0.770 | 0.984 | 0.063 | 0.352 | 0.084 |
| 2. Do you lack money to fulfill household needs? | 1.000 | 0.931 | 1.000 | 0.996 | 0.719 | 0.990 | 0.038 | 0.389 | 0.061 |
| 3. It is hard to make sure about access to different food types? | 1.000 | 0.988 | 1.000 | 0.976 | 0.865 | 0.992 | 0.094 | 0.232 | 0.053 |
| 4. Does anyone in your house use controlled medication? | 0.957 | 0.983 | 0.970 | 0.766 | 0.844 | 0.799 | 0.328 | 0.275 | 0.320 |
| 5. Does anyone in your house use 5, or more medications, on a daily basis? | 0.662 | 0.988 | 0.926 | 0.469 | 0.863 | 0.711 | 0.491 | 0.261 | 0.361 |
| 6. Does anyone in your house have a health condition that requires continuous care? | 0.927 | 0.968 | 0.963 | 0.711 | 0.795 | 0.782 | 0.449 | 0.383 | 0.374 |
| 7. Is anyone in your house impaired to perform daily activities? | 0.850 | 0.966 | 0.941 | 0.617 | 0.789 | 0.735 | 0.560 | 0.400 | 0.439 |
| 8. Does anyone in your house need help to accomplish its own daily healthcare procedures? | 0.742 | 0.961 | 0.887 | 0.525 | 0.777 | 0.657 | 0.538 | 0.378 | 0.478 |
| 9. Did anyone in your house have its mother absent in childhood? | 0.291 | 0.347 | 0.387 | 0.233 | 0.270 | 0.295 | 0.600 | 0.493 | 0.516 |
| 10. Did anyone in your house have an absent father in childhood? | 0.495 | 0.240 | 0.511 | 0.363 | 0.198 | 0.373 | 0.465 | 0.535 | 0.389 |
| 11. Has anyone in your house faced abandonment by the family? | 0.927 | 0.583 | 0.912 | 0.711 | 0.418 | 0.690 | 0.329 | 0.439 | 0.297 |
| 12. Does anyone in your house live close to violent people? | 0.868 | 0.936 | 0.488 | 0.636 | 0.727 | 0.358 | 0.481 | 0.115 | 0.616 |
| 13. Have anyone in your house been victim of violence? | 0.861 | 0.914 | 0.731 | 0.628 | 0.693 | 0.517 | 0.450 | 0.295 | 0.564 |
| 14. Is there violence in your house? | 0.914 | 0.963 | 0.349 | 0.692 | 0.782 | 0.272 | 0.381 | 0.021 | 0.503 |

I-UNICO, Unidimensional Congruence; I-ECV, Explained Common Variance; I-REAL, Residual Absolute Loadings; TrS, Training Sample; TsS, Test Sample; FS, Full Sample.

TrS primary data (Table 4) presented factorial data ranging from 0.597 to 0.975, communality ranging from 0.449 to 0.967, and Eta ranging from 0.640 to 0.941. The model recorded explained variable of 76.18%. Items 1 and 3 comprised dimension **<u>Income</u>**, items from 4 to 8 comprised dimension **<u>Healthcare</u>**, dimension **<u>Family</u>** was in items 9 to 11, and the single dimension called **<u>Violence</u>** was observed in items 12 to 14. Accordingly, the model points towards good factorial and interpretable (quantitatively) solution, with content alignment in coherent and interpretable (quantitatively) items. The final version of the scale represented reduction by approximately 63% in the 38 items assessed through judges’ panel in the first stage. This value is close to that presented by DeVellis [28], according to whom the researcher must project items lost by 50% throughout the process.

**Table 4.** Training dataset: Factorial loads, communality and Eta.

| Item | Factorial Load |  |  |  | h<sup>2</sup> | Pratt's Measure - (Eta) |  |  |  |
| --- | --- | --- | --- | --- | --- | --- | --- | --- | --- |
|  | Income | Healthcare | Family | Violence |  | Income | Healthcare | Family | Violence |
| 1. Is anyone in your house facing financial issues? | **0.893** | 0.027 | -0.012 | 0.059 | 0.849 | **0.904** | 0.098 | 0.000 | 0.146 |
| 2. Do you lack money to fulfill household needs? | **0.895** | -0.013 | 0.013 | -0.067 | 0.762 | **0.872** | 0.000 | 0.049 | 0.000 |
| 3. Is there any difficulty in making sure about food variety in your house? | **0.597** | 0.091 | 0.102 | 0.110 | 0.516 | **0.642** | 0.180 | 0.174 | 0.203 |
| 4. Does anyone in your house use controlled medication? | -0.100 | **0.693** | 0.101 | 0.067 | 0.500 | 0.000 | **0.680** | 0.146 | 0.130 |
| 5. Does anyone in your house use 5, or more, medications on a daily basis? | -0.080 | **0.711** | 0.031 | -0.118 | 0.449 | 0.000 | **0.668** | 0.051 | 0.000 |
| 6. Does anyone in your house have a health condition that requires continuous care? | -0.032 | **0.805** | 0.020 | 0.069 | 0.671 | 0.000 | **0.805** | 0.059 | 0.139 |
| 7. Is anyone in your house impaired to perform daily activities? | 0.094 | **0.862** | -0.131 | 0.029 | 0.798 | 0.189 | **0.869** | 0.000 | 0.084 |
| 8. Does anyone in your house need help to accomplish its own daily healthcare procedures? | 0.062 | **0.737** | -0.053 | -0.090 | 0.540 | 0.132 | **0.723** | 0.000 | 0.000 |
| 9. Did anyone in your house have its mother absent in childhood? | 0.012 | -0.140 | **0.798** | 0.013 | 0.630 | 0.042 | 0.000 | **0.790** | 0.061 |
| 10. Did anyone in your house have its father absent in childhood? | 0.056 | -0.033 | **0.741** | -0.097 | 0.514 | 0.099 | 0.000 | **0.710** | 0.000 |
| 11. Has anyone in your house faced abandonment by the family? | 0.018 | 0.148 | **0.630** | 0.073 | 0.504 | 0.065 | 0.203 | **0.658** | 0.163 |
| 12. Does anyone in your house live close to violent people? | 0.192 | -0.130 | -0.108 | **0.975** | 0.967 | 0.284 | 0.000 | 0.000 | **0.941** |
| 13. Have anyone in your house been victim of violence? | -0.118 | 0.086 | 0.290 | **0.605** | 0.574 | 0.000 | 0.146 | 0.379 | **0.640** |
| 14. Is there violence in your house? | -0.124 | 0.055 | -0.091 | **0.937** | 0.787 | 0.000 | 0.117 | 0.000 | **0.880** |

Based on the four dimensions composing the scale, it is possible taking into account the key role played by social determinants within the health/illness process. Dimensions embody items related to income, social and family cohesion, and to life and housing conditions associated with psychosocial and behavioral aspects [62]. Thus, it extrapolates the biological view of health, which is overall acknowledged in a reductionist way, centered in medical practices [62-65]. Nevertheless, the present instrument emerges as multi-disciplinary work-tool available for PHC teams that have the potential to promote social justice by taking into consideration social inequities at the time to plan healthcare services.

Factorial loads recorded for the TsS dataset (Table 5) ranged from 0.643 to 0.976, communality ranged from 0.482 to 0.910 and Eta ranged from 0.658 to 0.951. This model presented explained variance of 76.18%. Once again, the observed model was equal to the training dataset model; consequently, it was quantitatively and qualitatively interpretable.

**Table 5.** Test dataset: Factor loading, communality and Eta.

| Item | Factorial Load |  |  |  | h<sup>2</sup> | Pratt's Measure - (Eta) |  |  |  |
| --- | --- | --- | --- | --- | --- | --- | --- | --- | --- |
|  | Income | Healthcare | Family | Violence |  | Income | Healthcare | Family | Violence |
| 1. Is anyone in your house facing financial issues? | **0.801** | 0.032 | 0.046 | 0.056 | 0.702 | **0.818** | 0.108 | 0.128 | 0.075 |
| 2. Do you lack money to fulfill household needs? | **0.976** | -0.072 | 0.013 | -0.008 | 0.910 | **0.951** | 0.000 | 0.068 | 0.000 |
| 3. Is there any difficulty in making sure about food variety in your house? | **0.805** | 0.063 | -0.084 | -0.006 | 0.647 | **0.790** | 0.151 | 0.000 | 0.000 |
| 4. Does anyone in your house use controlled medication? | 0.059 | **0.671** | -0.009 | -0.006 | 0.482 | 0.138 | **0.681** | 0.000 | 0.000 |
| 5. Does anyone in your house use 5, or more, medications on a daily basis? | 0.141 | **0.639** | -0.078 | 0.066 | 0.493 | 0.227 | **0.658** | 0.000 | 0.094 |
| 6. Does anyone in your house have a health condition that requires continuous care? | -0.047 | **0.882** | 0.023 | -0.012 | 0.753 | 0.000 | **0.865** | 0.063 | 0.000 |
| 7. Is anyone in your house impaired to perform daily activities? | 0.001 | **0.881** | -0.030 | 0.011 | 0.770 | 0.015 | **0.876** | 0.000 | 0.035 |
| 8. Does anyone in your house need help to accomplish its own daily healthcare procedures? | -0.080 | **0.856** | 0.037 | -0.033 | 0.690 | 0.000 | **0.827** | 0.078 | 0.000 |
| 9. Did anyone in your house have its mother absent in childhood? | -0.024 | -0.030 | **0.727** | 0.036 | 0.514 | 0.000 | 0.000 | **0.715** | 0.059 |
| 10. Did anyone in your house have its father absent in childhood? | -0.015 | -0.090 | **0.752** | 0.037 | 0.546 | 0.000 | 0.000 | **0.736** | 0.059 |
| 11. Has anyone in your house faced abandonment by the family? | -0.035 | 0.077 | **0.757** | -0.062 | 0.575 | 0.000 | 0.123 | **0.748** | 0.000 |
| 12. Does anyone in your house live close to violent people? | 0.067 | -0.016 | -0.037 | **0.918** | 0.843 | 0.076 | 0.000 | 0.000 | **0.915** |
| 13. Have anyone in your house been victim of violence? | 0.138 | 0.096 | 0.310 | **0.643** | 0.652 | 0.211 | 0.164 | 0.370 | **0.666** |
| 14. Is there violence in your house? | -0.177 | -0.041 | -0.172 | **0.923** | 0.893 | 0.196 | 0.042 | 0.167 | **0.909** |

**Table 6.** Full dataset: Factor loading, communality and Eta.

| Item | Factorial load |  |  |  | h<sup>2</sup> | Pratt's Measure - (Eta) |  |  |  |
| --- | --- | --- | --- | --- | --- | --- | --- | --- | --- |
|  | Income | Healthcare | Family | Violence |  | Income | Healthcare | Family | Violence |
| 1. Is anyone in your house facing financial issues? | **0.733** | 0.009 | 0.004 | 0.023 | 0.548 | **0.736** | 0.049 | 0.029 | 0.047 |
| 2. Do you lack money to fulfill household needs? | **0.785** | -0.045 | -0.015 | -0.020 | 0.585 | **0.765** | 0.000 | 0.000 | 0.000 |
| 3. Is there any difficulty in making sure about food variety in your house? | **0.555** | 0.059 | 0.019 | 0.027 | 0.346 | **0.569** | 0.124 | 0.060 | 0.050 |
| 4. Does anyone in your house use controlled medication? | -0.001 | **0.523** | 0.056 | 0.016 | 0.287 | 0.000 | **0.528** | 0.087 | 0.035 |
| 5. Does anyone in your house use 5, or more, medications a day? | 0.019 | **0.489** | -0.012 | -0.010 | 0.243 | 0.060 | **0.489** | 0.000 | 0.000 |
| 6. Does anyone in your house have a health condition that requires continuous care? | 0.007 | **0.639** | 0.032 | 0.013 | 0.421 | 0.041 | **0.644** | 0.065 | 0.032 |
| 7. Is anyone in your house impaired to perform daily activities? | 0.008 | **0.723** | -0.048 | 0.001 | 0.518 | 0.044 | **0.718** | 0.000 | 0.008 |
| 8. Does anyone in your house need help to accomplish its own daily healthcare procedures? | -0.036 | **0.624** | -0.017 | -0.024 | 0.371 | 0.000 | **0.609** | 0.000 | 0.000 |
| 9. Did anyone in your house have its mother absent in childhood? | -0.027 | -0.049 | **0.594** | 0.004 | 0.339 | 0.000 | 0.000 | **0.582** | 0.013 |
| 10. Did anyone in your house have an absent father in childhood? | 0.009 | -0.043 | **0.552** | -0.009 | 0.302 | 0.037 | 0.000 | **0.548** | 0.000 |
| 11. Has anyone in your house faced abandonment by the family? | 0.004 | 0.060 | **0.471** | 0.001 | 0.236 | 0.026 | 0.089 | **0.476** | 0.007 |
| 12. Does anyone in your house live close to violent people? | 0.025 | -0.034 | 0.011 | **0.743** | 0.554 | 0.047 | 0.000 | 0.029 | **0.742** |
| 13. Has anyone in your house been victim of violence? | 0.028 | 0.084 | 0.273 | **0.308** | 0.212 | 0.069 | 0.119 | 0.296 | **0.325** |
| 14. Is there violence in your house? | -0.047 | -0.005 | -0.018 | **0.597** | 0.351 | 0.000 | 0.000 | 0.000 | **0.593** |

These results ranged from 0.308 to 0.785 for factorial loads, from 0.212 to 0.967 for communality, and Eta ranged from 0.640 to 0.941. The model based on the total sample recorded explained variance of 79.02%. Number of dimensions’ stability and model interpretability are essential aspects of dimensionality. It reinforces the relevance of carrying out the cross-validation.

Reliability indices between analysis datasets ranged from 0.69 to 0.71 in Cronbach’s alpha, it reached 0.70 in the three dataset for Omega, it ranged from 0.83 to 0.84 for glb and from 0.80 to 0.96 ORION, between dimensions and datasets. Factorial solution quality indices also showed adequate levels, and this finding reinforced the model’s stability (Table 7). Accordingly, the set of applied techniques and indices pointed towards a set of internal structure validity evidences that are adequate, consistent, robust and interpretable.

**Table 7.**
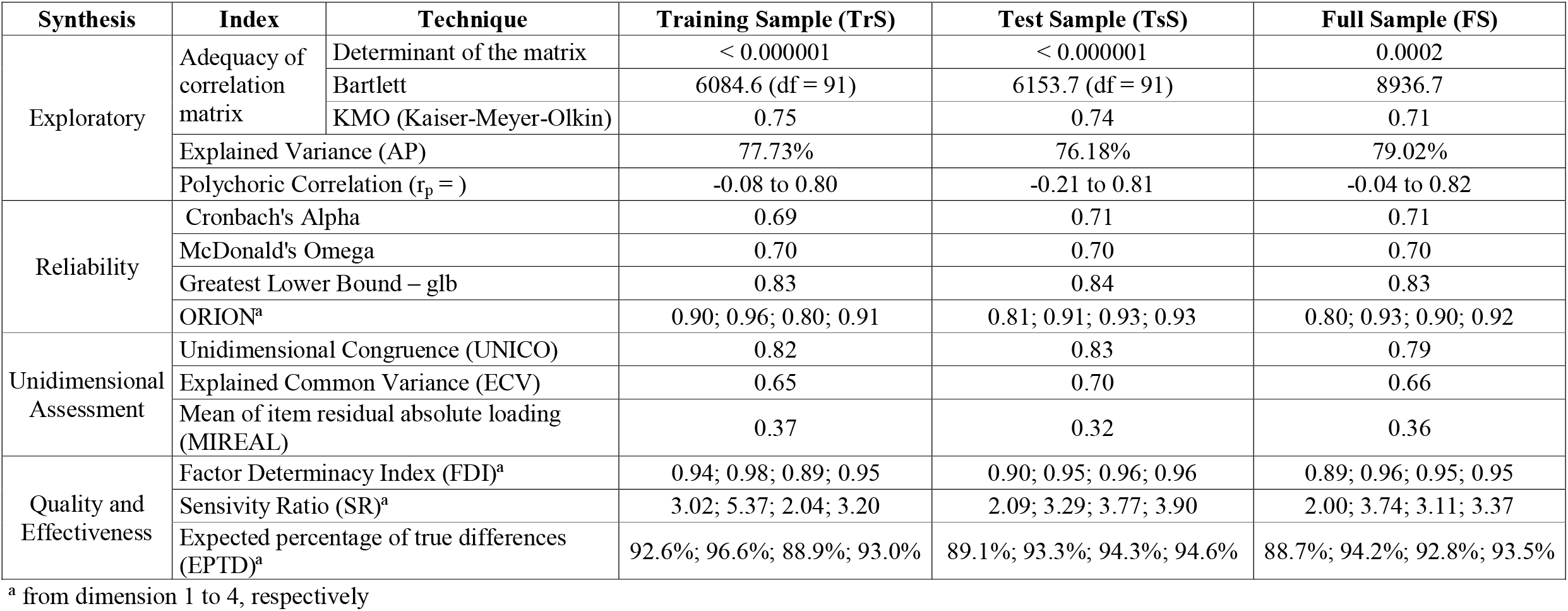
Synthesis of the models.

These results ranged from 0.308 to 0.785 for factorial loads, from 0.212 to 0.967 for communality, and Eta ranged from 0.640 to 0.941. The model based on the total sample recorded explained variance of 79.02%. Number of dimensions’ stability and model interpretability are essential aspects of dimensionality. It reinforces the relevance of carrying out the cross-validation.

Reliability indices between analysis datasets ranged from 0.69 to 0.71 in Cronbach’s alpha, it reached 0.70 in the three dataset for Omega, it ranged from 0.83 to 0.84 for glb and from 0.80 to 0.96 ORION, between dimensions and datasets. Factorial solution quality indices also showed adequate levels, and this finding reinforced the model’s stability (Table 7). Accordingly, the set of applied techniques and indices pointed towards a set of internal structure validity evidences that are adequate, consistent, robust and interpretable.

#### Standardization

The internal structure of the instrument was extrapolated and found. Now, the descriptive study and score standardization will be addressed to allow instrument interpretability and participants’ proper classification based on the scores. Accordingly, Table 8 depicts the frequency of answers to items in the questionnaire. There was clear prevalence of “No” answers for all items in the instrument. Some items presented higher frequency of “yes” answers: “Lack of money to fulfill household needs” (40.34%), “someone in the house uses controlled medication” (36.41%), “someone in the house has a health condition that requires continuous care” (36.77%) and “someone in the house had an absent father in childhood” (34.07%).

**Table 8.** Frequency of 397 answers for the items.

| Item | Frequency of answer to the item (N/%) |  |  |
| --- | --- | --- | --- |
|  | No | Yes | Missing |
| 1. Is anyone in your house facing financial issues? | 870 (69.10) | 377 (29.94) | 12 (0.95) |
| 2. Do you lack money to fulfill household needs? | 742 (58.93) | 508 (40.34) | 9 (0.71) |
| 3. Is there any difficulty in making sure about food variety in your house? | 928 (73.70) | 322 (25.57) | 9 (0.71) |
| 4. Does anyone in your house use controlled medication? | 793 (62.98) | 461 (36.61) | 5 (0.39) |
| 5. Does anyone in your house use 5, or more, medications a day? | 1,020 (81.01) | 231 (18.34) | 8 (0.63) |
| 6. Does anyone in your house have a health condition that requires continuous care? | 788 (62.58) | 463 (36.77) | 8 (0.63) |
| 7. Is anyone in your house impaired to perform daily activities? | 1,026 (81.43) | 232 (18.42) | 1 (0.07) |
| 8. Does anyone in your house need help to accomplish its own daily healthcare procedures? | 1,066 (84.67) | 190 (15.09) | 3 (0.23) |
| 9. Did anyone in your house have its mother absent in childhood? | 1,048 (83.24) | 207 (16.44) | 4 (0.31) |
| 10. Did anyone in your house have an absent father in childhood? | 826 (65.60) | 429 (34.07) | 4 (0.31) |
| 11. Has anyone in your house faced abandonment by the family? | 1,129 (89.67) | 125 (9.92) | 5 (0.39) |
| 12. Does anyone in your house live close to violent people? | 1,221 (96.98) | 34 (2.70) | 4 (0.31) |
| 13. Has anyone in your house been victim of violence? | 1,088 (86.41) | 167 (13.26) | 4 (0.31) |
| 14. Is there violence in your house? | 1,232 (97.85) | 25 (1.98) | 2 (0.15) |

Table 9 presents the scores recorded for the dimensions and the general score of the Family Vulnerability Scale. All dimensions had all their amplitudes answered. Dimensions Income, Family and Violence recorded median = 0, Healthcare showed median = 1 and total score recorded median = 2. An interesting aspect of the total score lies on the fact that amplitude ranged from 0 to 14 and the maximum score recorded in the current sample reached 12. Medians in the minimum limit, and close to it, previously pointed out that the instrument can accurately differentiate individuals who are eventually facing family vulnerability situations.

**Table 9.**
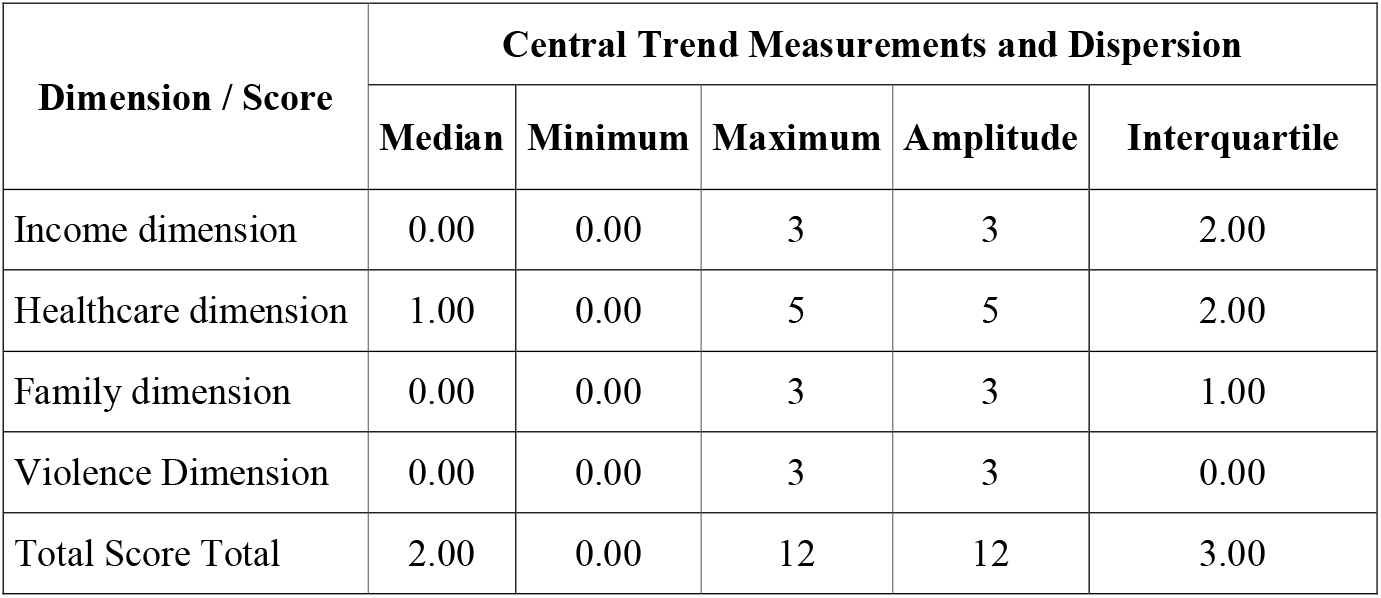
Description of dimensions and scores recorded for the Family Vulnerability Scale.

Because these scores are closer to the minimum limit, they only started presenting greater difference when they got far from the median that, in this case, was close to the minimum; therefore, in the upper quartile. Thus, three initial classifications were suggested: model 1 had cut in the median (low and high vulnerability), model 2 scores were separated until percentile 75, from 76 to 89, and higher than 90; model 3 scores were separated up to percentile 70, from 70 to 89, or higher.

The discriminant analysis of each classification developed to assess whether it was possible accurately identifying participants within the limit was applied after the first cuts were made.

The first analysis adopted a binary classification (low and high). The discriminant analysis showed MBox = 446.58 p < 0.001. λ_wilks_ = 0.36; F_(1, 1257)_ = 1,268.77; p < 0.001; canonical correlation = 0.797; the model with two limits properly classified 85.7% of the cases. The discriminant analysis applied to model 2 was MBox = 49.64 p < 0.001. λ_wilks_ = 0.18; F_(2, 1256)_ = 2,094.25; p < 0.001; canonical correlation = 0.907. Model 2 properly classified 100% of cases. The analysis applied to model 3 presented MBox = 49.64 p < 0.001. λ_wilks_ = 0.25; F_(2, 1256)_ = 1,838.71; p < 0.001; canonical correlation = 0.863; it was possible properly classifying 89% of cases. The recommended classification and scores interpretations are depicted in Table 10.

**Table 10.** Limits, classification and interpretation of Family Vulnerability Scale scores.

| Classification results | Percentile | Name | Score |
| --- | --- | --- | --- |
| Limit | Up to 75 | low | 0 to 4 |
|  | 76 to 89 | Moderate | 5 to 6 |
|  | Higher than 90 | High | Higher than 7 |

### Family Vulnerability Scale – final version

The final version of the Family Vulnerability scale (EVFAM-BR) comprised 14 items (or questions) applied to each family in the PHC territory in Brazil, due to the action by Community Health Agents (CHA). EVFAM-BR has four dimensions; each one of them has a score corresponding to the number of items in the dimension – at the end of its application, the score must range from zero (0) to fourteen (14). Healthcare is the dimension accounting for the largest number of items; consequently, it has the greatest potential in the scale (n=5). The final EVFAM-BR presented three family vulnerability classification limits if one sums the scores of each dimension: Low (0 to 4), Moderate (5 to 6) and High (7 to 14). Table 11 presents a summary of the Family Vulnerability Scale, and its respective dimensions, items and scores.

**Table 11.** Family Vulnerability Scale (EVFAM-BR).

| **Dimension** | **Item** | **Item score** | **Dimension score** |
| --- | --- | --- | --- |
| Income | 1. Is anyone in your house facing financial issues? | 1 | 3 |
|  | 2. Do you lack money to fulfill household needs? | 1 |  |
|  | 3. Is there any difficulty in making sure about food variety in your house? | 1 |  |
| Healthcare | 4. Does anyone in your house use controlled medication? | 1 | 5 |
|  | 5. Does anyone in your house use 5, or more, medications on a daily basis? | 1 |  |
|  | 6. Does anyone in your house have a health condition that requires continuous care? | 1 |  |
|  | 7. Is anyone in your house impaired to perform daily activities? | 1 |  |
|  | 8. Does anyone in your house need help to accomplish its own daily healthcare procedures? | 1 |  |
| Family | 9. Did anyone in your house have its mother absent in childhood? | 1 | 3 |
|  | 10. Did anyone in your house have an absent father in childhood? | 1 |  |
|  | 11. Has anyone in your house faced abandonment by the family? | 1 |  |
| Violence | 12. Does anyone in your house live close to violent people? | 1 | 3 |
|  | 13. Has anyone in your house been victim of violence? | 1 |  |
|  | 14. Is there violence in your house? | 1 |  |
| **Total** |  | **14** | **14** |

## Discussion

The Family Vulnerability Scale (EVFAM-BR) has shown evidences of content and internal structure validity based on the multi-regional and multi-professional context, and this finding allows measuring family vulnerability in Brazil.

EVFAM-BR comprises 14 items distributed into the following dimensions: Income, Healthcare, Family and Violence. It is worth highlighting that EVFAM-BR aims at measuring social vulnerability within the family context; consequently, all items refer to the family nucleus, they are not oriented to one specific resident, or to the respondent, itself.

Just to exemplify EVFAM-BR application to a family in a given local or time: one of the residents in a given house is facing financial issues. However, lack of money to fulfill household needs is not identified and there is no hard time accessing different food types; one of the residents has a chronic disease that requires continuous care and uses controlled medication (less than 5 medication types a day), but none of the residents has any difficulty in performing daily activities and does not need daily healthcare; none of the residents lacked mother or father presence in childhood and no family member faced abandonment situations; there was no violence in the house and no one in the house lives with violent people, but one of the residents was a victim of violence. Given the positive answers to items financial issue by one of the residents (1), health condition requiring continuous care (1), the use of medication (1) and person who was violence victim (1), this family would reach score 4, and – based on the classification limit of EVFAM-BR final score - it represents a family classified as “low family vulnerability”.

The literature consistently states that income is a relevant social health determinant and that it must be taken into account to allow planning equitable healthcare provision [66]. The subsequent discussion about this topic led to different views on how income inequality affects health. Rich or poor individuals, in low social cohesion societies, would be the target of problems such as crime, lack of public investments, and it makes people adopt unhealthy behaviors such as smoking, excessive alcohol consumption and sedentary life [67]. These outcomes can help better understanding the relationship between variables linked to income and enable interventions at macroeconomic level, as well as assessing these changes in population health.

The healthcare dimension is timely, given the accelerated population aging in the country and abroad, a fact that demands healthcare services’ reorganization to continuously fulfill population needs, in an organized way, based on quality and safety. With respect to the family dimension, several studies have shown the association between absence of parents and different health outcomes, among them one finds cognitive development loss [68,69], impacts on mental health [70,71], and early development of risk behavior for health, such as smoking and alcohol abuse [72].

Finally, dimension “violence” corroborated the discussion observed in WHO’s 2030 agenda for the sustainable development of millennium goals; this agenda highlights violence prevention as fundamental component for both development and improved quality of life, worldwide. It is known that violence affects health and broadens the demands for healthcare in a way wider than simply through initial trauma, it extrapolates the probability of other important causes for diseases and death [73]. It is possible identifying association among exposure to violence, undesired health outcomes and unhealthy behaviors, such as drug and alcohol abuse, mainly among low-income mothers in urban locations [74].

Although the urgency in having research focused on low and medium income countries that account for 90% of the global violence, only 10% of studies in this field are performed in them [73]. Thus, EVFAM-BR emerges as a tool to allow structurally and routinely introducing the approach of social factors associated with the health/illness process in healthcare services, in developing countries.

Accordingly, EVFARM-BR validity evidences, along with the four family vulnerability strata proposed based on its application, present the potential of this tool to help the role played by PHCs in performing their attributes, mainly in coordinating caregiving based on population-base management within the community and family context [75].

It is important highlighting that EVFAM-BR is an instrument presenting robust and synthesized evidences that, at first, demand low workload investment by professionals and low financial resources. Because it is an objective instrument (only “yes” and “no” answers are expected), it suggests that all PHC professionals must be trained to use it. This instrument emerges as powerful tool to support the work by community health agents (CHA), since these actors are community members and are closely bond to families in the territory; this process makes the “interview environment” more comfortable and trustful for users who answer the questions on behalf of the household. Thus, EVFAM-BR can be applied through printed materials or, yet, online, since it is a tool used as work instrument added to the teams’ routine. One must take into account its potential for inclusion in the digital registration system of the Brazilian Unified Health System (SUS).

It is possible guiding the teams at the time to plan their actions and health interventions (based on exposing families to conditions that increase vulnerability and risk to develop illnesses) by applying EVFAM-BR and by interpreting its household-classification results based on the four predicted strata. It is worth highlighting how vulnerability contexts can be changed overtime; instrument application must be periodical to keep teams’ planning updated, according to population needs.

Among limitations of the current study, one finds sampling based on convenience. It does not ensure statistical results’ reliability. However, the study was carried out in different socioeconomic, demographic and cultural contexts, since it encompassed participants from the five geographic regions in Brazil. Yet, it is important pointing out the potential of carrying out research in the PHC context in order to allow the participation of people with different demands, needs and life conditions, who seek care in this service.

Highlights in the present research are data collection by professionals outside the assessed services who were trained to carry out the interviews, as well as the use of robust techniques to identify EVFAM-BR validity evidences; among them, CVR presents a sophisticated and more adequate method [76] in comparison to the proposed alternatives [77]. CVR calculation takes into consideration the number of judges [24,25] and it minimizes the increase in random compliance [24], a fact that allows adopting a large number of judges to judge the instrument. The adoption of a multi-disciplinary panel of judges comprising researchers, translators, health professionals, methodology experts and lay people leads to more consistent results from the judges’ panel [77-79].

It is important pinpointing the need of implementing research in order to identify the potential and challenges of using EVFAM-BR in the routine of PCH’s services in different Brazilian contexts and, yet, in countries presenting similar health system features, as well as population socioeconomic, demographic, sanitary and epidemiological features.

## Conclusion

Given the set of herein employed techniques, it is possible stating that the set of content validity and EVFAM-BR internal structure evidences are adequate, consistent, reliable and robust, as well as that the cross-validation method ensured model reliability and replicability. A synthetic scale was presented, and it is capable of accurately measuring and differentiating familiar vulnerability.

## Data Availability

All relevant data are within the manuscript and its Supporting Information files.

