## Supplemental material for "Family Vulnerability Scale: evidence of content and internal structure validity"

| **Dimension** | **Item** | **Item score**  **1 (Yes) or 0 (No)** |
| --- | --- | --- |
| Income | 1. Is anyone in your house facing financial issues? |  |
|  | 2. Do you lack money to fulfill household needs? |  |
|  | 3. Is there any difficulty in making sure about food variety in your house? |  |
| Healthcare | 4. Does anyone in your house use controlled medication? |  |
|  | 5. Does anyone in your house use 5, or more, medications on a daily basis? |  |
|  | 6. Does anyone in your house have a health condition that requires continuous care? |  |
|  | 7. Is anyone in your house impaired to perform daily activities? |  |
|  | 8. Does anyone in your house need help to accomplish its own daily healthcare procedures? |  |
| Family | 9. Did anyone in your house have its mother absent in childhood? |  |
|  | 10. Did anyone in your house have an absent father in childhood? |  |
|  | 11. Has anyone in your house faced abandonment by the family? |  |
| Violence | 12. Does anyone in your house live close to violent people? |  |
|  | 13. Has anyone in your house been victim of violence? |  |
|  | 14. Is there violence in your house? |  |
| **Total** | |  |

**Supporting Information 1. Family Vulnerability Scale (EVFAM-BR).**
